# Can Psychedelic Use Benefit Meditation Practice? Examining Individual, Psychedelic, and Meditation-Related Factors

**DOI:** 10.1101/2024.08.27.24312677

**Authors:** Zishan Jiwani, Simon B. Goldberg, Jack Stroud, Jacob Young, John Curtin, John D. Dunne, Otto Simonsson, Christian A. Webb, Robin Carhart-Harris, Marco Schlosser

## Abstract

**Introduction:** Meditation practice and psychedelic use have attracted increasing attention in the public sphere and scientific research. Both methods induce non-ordinary states of consciousness that may have significant therapeutic benefits. Thus, there is growing scientific interest in potential synergies between psychedelic use and meditation practice with some research suggesting that psychedelics may benefit meditation practice. The present study examined individual, psychedelic-related, and meditation-related factors to determine under what conditions meditators perceive psychedelic use as beneficial for their meditation practice.

**Method:** Participants (*N* = 863) who had reported psychedelic use and a regular meditation practice (at least 3 times per week during the last 12 months) were included in the study. To accommodate a large number of variables, machine learning (i.e., elastic net, random forest) was used to analyze the data.

**Results:** Most participants (*n* = 634, 73.5%) found psychedelic use to have a positive influence on their quality of meditation. Twenty-eight variables showed significant zero-order associations with perceived benefits even following a correction. Elastic net had the best performance (R^2^ = .266) and was used to identify the most important features. Across 53 variables, the model found that greater use of psychedelics, intention setting during psychedelic use, agreeableness, and exposure to N,N-Dimethyltryptamine (N,N-DMT) were most likely to be associated with the perception that psychedelics benefit meditation practice. The results were consistent across several different approaches used to identify the most important variables (i.e., Shapley values, feature ablation).

**Discussion:** Results suggest that most meditators found psychedelic use to have a positive influence on their meditation practice, with: 1) regularity of psychedelic use, 2) the setting of intentions for psychedelic use, 3) having an agreeable personality, and 4) reported use of N,N-DMT being the most likely predictors of perceiving psychedelic use as beneficial. Longitudinal designs and randomized trials manipulating psychedelic use are needed to establish causality.

---

Meditation practices and psychedelic use have both received increased research and popular interest in the past two decades. As methods for inducing non-ordinary states of consciousness, meditation practices, and psychedelic use may have therapeutic benefits, with a “psychedelic renaissance” in particular becoming highly visible in the past several years (1–7). In terms of prevalence, a population-based survey conducted by the Centers for Disease Control in the United States (US) suggested that in 2022, 17.3% of Americans engaged in some form of meditation practice (8). Similarly, data from the National Survey on Drug Use and Health conducted in 2018 suggested that approximately 15.9% of the US population had lifetime exposure to psychedelics (9). Research on the therapeutic effects of meditation practices, while well established, is still ongoing, and in the case of psychedelic use, it is nascent. Nevertheless, recent systematic reviews and meta-analyses suggest that both meditation practice and psychedelic use may be beneficial for a range of psychological symptoms (10–14).

There is growing scientific interest in exploring potential synergies between psychedelic use and meditation practices (4,15–21). Psychedelics and at least some forms of meditation may target similar brain regions, have some degree of phenomenological overlap and may have shared or complementary psychological mechanisms of change (22). For instance, mindfulness-style meditation practices and psychedelic use have been associated with changes in the Default Mode Network (DMN), a network of interacting regions of the brain that are activated when a person is not focused on the external world (23,24). Changes in the DMN induced by psychedelics or by mindfulness-style meditation have been associated with experiences of a loss of subjective identity, also known as “ego dissolution” which has been found to be associated with long-term wellbeing (25,26). Additionally, both practices may also share psychological mechanisms such as acceptance, decentering, and cognitive flexibility (16,25).

Although the literature is still quite small, there is some evidence that psychedelic use may support meditation practices. In a recent experimental study, psilocybin or placebo was given to 39 meditators during a five-day mindfulness meditation retreat (21,27,28). Participants who received psilocybin experienced greater ego dissolution during the retreat and reported larger positive changes in psychosocial functioning at a 4-month follow-up, suggesting a synergistic relationship between simultaneous psychedelic use and mindfulness meditation practice (i.e., using psychedelics while or just before meditating). Additionally, a large cross-sectional study (*n* = 2,822) examining psychedelic use and meditation practice (i.e., using both psychedelics and meditation but not necessarily at the same time) found that psychedelic use was associated with a higher degree of current mindfulness meditation practice (20). A recent longitudinal replication (*n* = 9732) confirmed these findings (29). Finally, a recent qualitative study examined written accounts of participants who had combined psychedelic use with meditation practices outside of treatment, retreat, or research settings. Findings from this online study suggested that most participants perceived that simultaneous use enhanced either their meditation practice, psychedelic experience, or both (30). Many participants also found subjective similarities between meditation practice and psychedelic use but reported that the intensity of mystical experience was greater with psychedelic use.

One potential reason that psychedelics may positively enhance the potential impact of meditation practice on psychological outcomes is that psychedelic use itself may increase one’s levels of mindfulness (12,18,31). Mindfulness is a semantically ambiguous construct with a rich conceptual history (32–36). Although contemplative scientists and scholars have not agreed upon a universally accepted definition of mindfulness or its constituents (see Van Dam et al., 2018), the scientific literature generally cites Jon Kabat-Zinn’s operational definition: ‘paying attention in a particular way: on purpose, in the present moment, and non-judgmentally’ (38,39).

Mindfulness and related attentional skills, such as decentering (i.e., the capacity to step outside the immediate experience) have also been defined as a form of heightened and malleable attentiveness to one’s experience including thoughts, feelings, bodily sensations, and perceptual impressions (40–43).

Several studies have investigated mindfulness in the context of psychedelic use. For instance, there is some indication that following psychedelic use, participants may experience greater mindfulness and related capacities including decentering for weeks or even months following initial use (12,44–48). Furthermore, psychedelic-induced mystical experiences which occasion changes in perception and cognition share similarities to experiences associated with advanced meditative states (26,49). Thus, it is possible that for meditators, psychedelic-induced experiences in supportive settings may make evident previously unexperienced aspects of human potential that may motivate their desire to deepen their understanding of such experiences through ongoing meditation practice.

Despite the potential synergy between psychedelic use and meditation practice, it has not been established to what degree meditators report benefits to their meditation associated with their use of psychedelics. In theory, there are a range of factors that may influence the degree to which meditators perceive benefits from psychedelic use. One category of factors is associated with the psychedelic itself (i.e., psychedelic-related factors). It has long been recognized that both the short-term and long-term effects of psychedelic use are dependent on set and setting (50). Set describes psychological, social, and cultural factors such as personality, preparation, expectation, intention, and worldview; setting captures the physical and sociocultural environment in which the experience unfolds (51,52). In addition, psychedelic type, dose, and frequency of use may also impact the psychedelic experience and its after-effects in ways that may differentially influence meditation practice (31).

A second set of factors relates to the meditators’ meditation practice (i.e., meditation-related factors). Meditation is a multidimensional construct comprising a wide range of distinct practices that can be cultivated to differing depths of subtlety and meditative skill (42,53,54). Given this complexity and the absence of a single unifying framework of meditation practices that could be used to consistently implement practice paradigms across scientific studies, meditation research has frequently narrowed its investigation to a subset of meditation practices, such as mindfulness practices. Naturally, a large part of the existing research at the intersection of psychedelic use and meditation practice has also reflected this mindfulness-centered focus.

Refining our understanding of the potential synergy between psychedelic use and meditation practice will require a more nuanced delineation and examination of meditation practices – including practices that, until recently, have not received scientific attention (e.g., meditative absorptions [Pali: *jhānas*]; 55–57) – as well as their purported mechanisms of action (see e.g., Lutz et al., 2015; Schlosser et al., 2022; Simonds, 2023). Furthermore, it may also be important whether an individual engages in psychedelic use prior to starting a meditation practice or vice versa.

Finally, a third set of factors relates to the meditators themselves (i.e., individual factors).

For instance, personality factors such as openness to experience and agreeableness have been associated with mystical-type experiences for psychedelic users (61,62). Demographic characteristics, such as gender, have also been found to be associated with psychedelic use in a community sample (63) and may influence the perceived benefit of psychedelics on meditation practice.

## Aims of the Present Study

While psychedelics may benefit meditation practice, it is unclear which factors might support the perceived benefits of psychedelic use. Using a large sample of meditators with exposure to psychedelics, the present study examined a range of factors, broadly categorized as individual, psychedelic-related, and meditation-related factors, to assess under which conditions a meditator might perceive their psychedelic use as beneficial for their meditation practice. To include a large range of potential factors, we used machine learning methods which are better able to handle the inclusion of a large number of predictor variables (including correlated variables) than traditional regression approaches (64,65). Overall, we intended to contribute to this nascent research area by including (i) a multidimensional conceptualization and assessment of meditation practices and meditation-related factors, (ii) a comprehensive assessment of psychedelic-related factors (including set and setting of naturalistic use), and, (iii) given the absence of standardized self-report measures for capturing the perceived effects of psychedelics on meditation practice, tailor-made questions that examine these relationships and provide a basis for future research in this area.

## Method

### Participants

A convenience sample was recruited through social media platforms (e.g., Twitter, Reddit, Facebook) and targeted advertising to meditation teachers, contemplative communities, meditation centers, and mindfulness associations. Any adult (≥ 18 years old) was eligible if they self-reported a good understanding of English and a regular meditation practice (at least 3 times per week during the last 12 months). While participants were told in the recruitment materials that the study aimed to understand the relationship between psychedelics and meditation, exposure to psychedelics was not required for participation. A total of 1049 individuals participated in the study. However, only participants who had both meditation and psychedelic exposure (*N* = 863) were included in the current analyses. The average participant age was 37.7 years (*SD* = 12.6) and ranged from 18-81 years. The sample had a larger percentage of participants who were assigned male sex at birth (*n* = 682, 79.4%) and reported on average 17.5 years of education (*SD* = 3.3). See Table 1 for sample demographics.

**Table 1.**
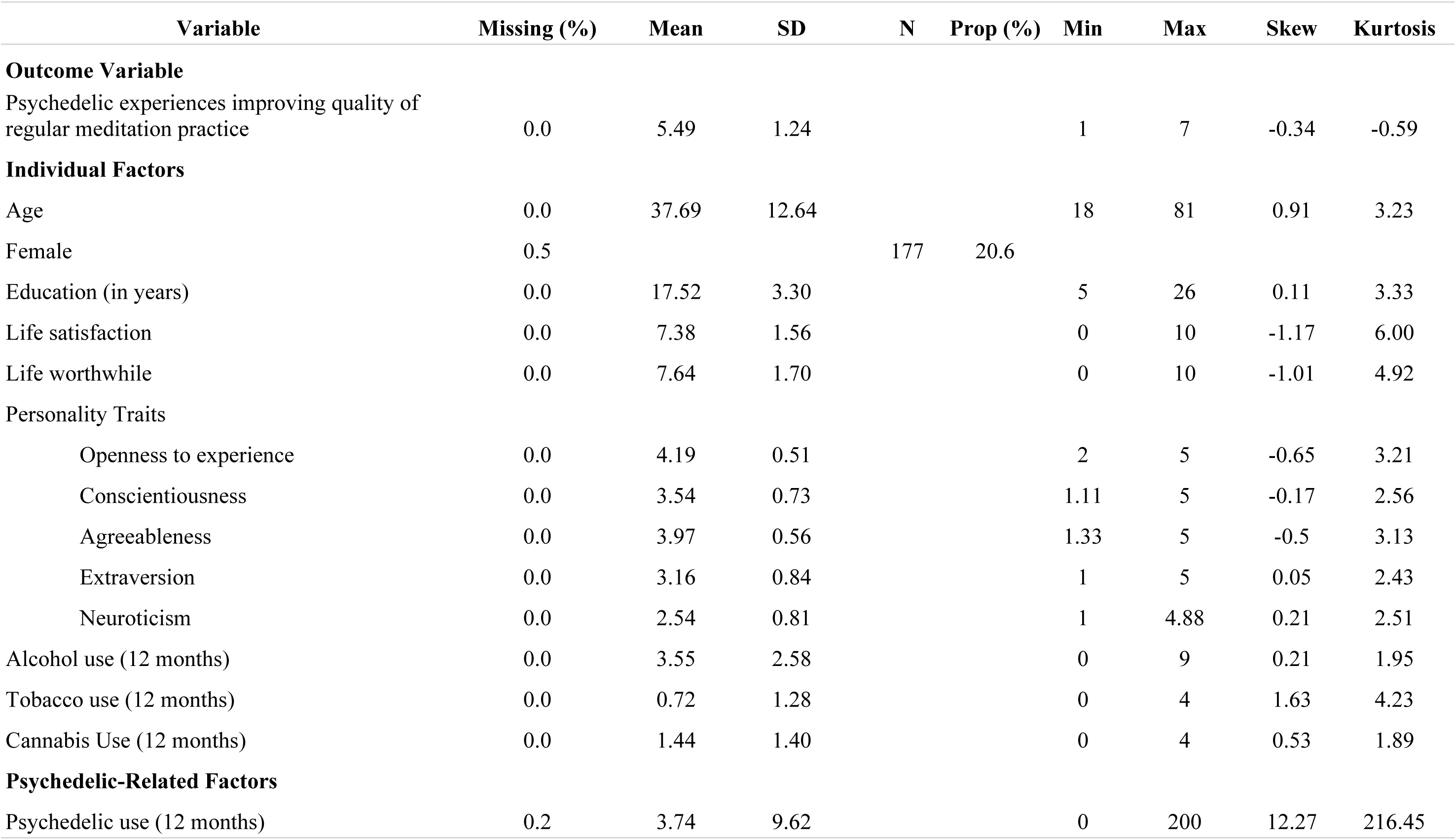
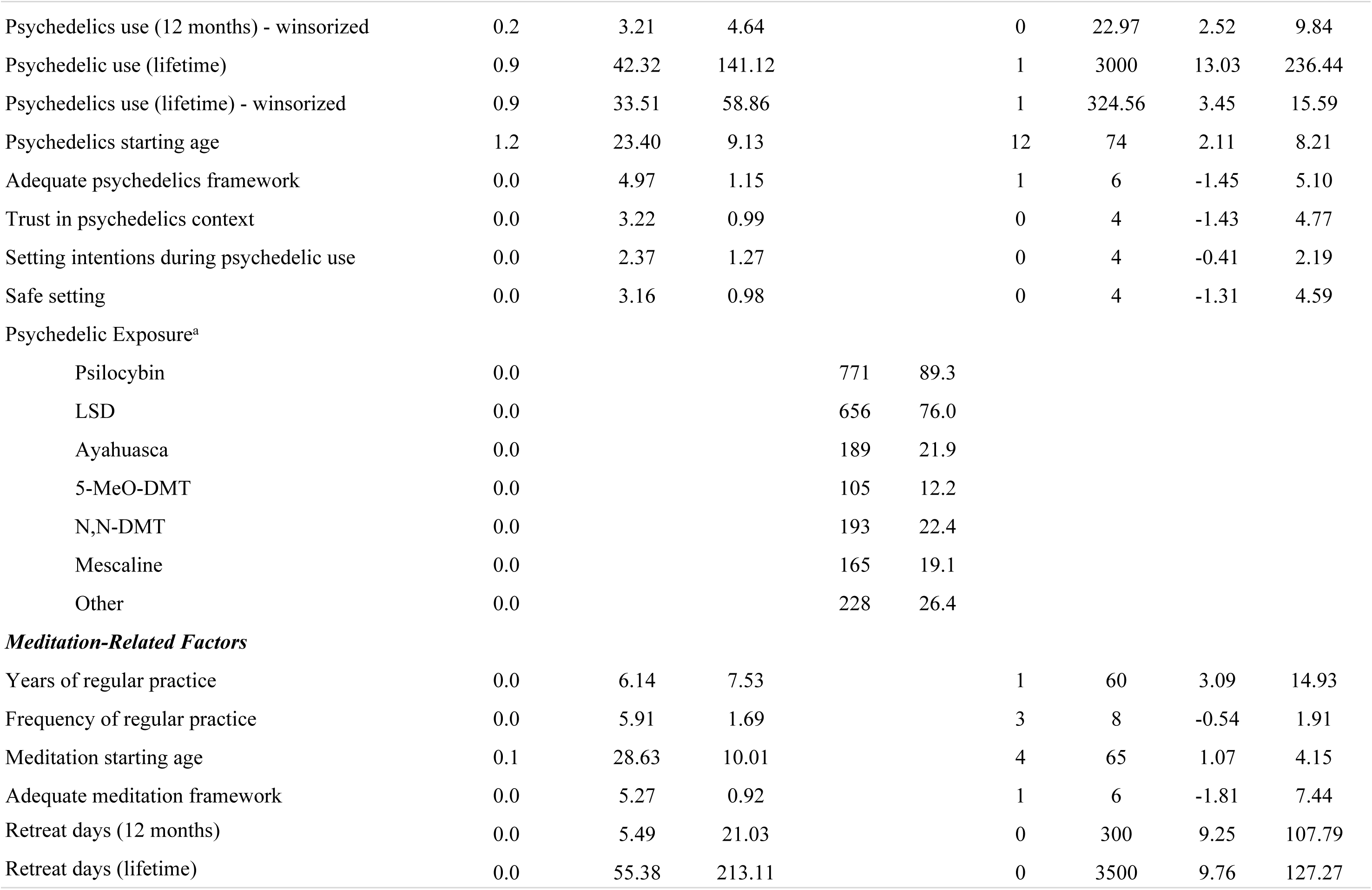
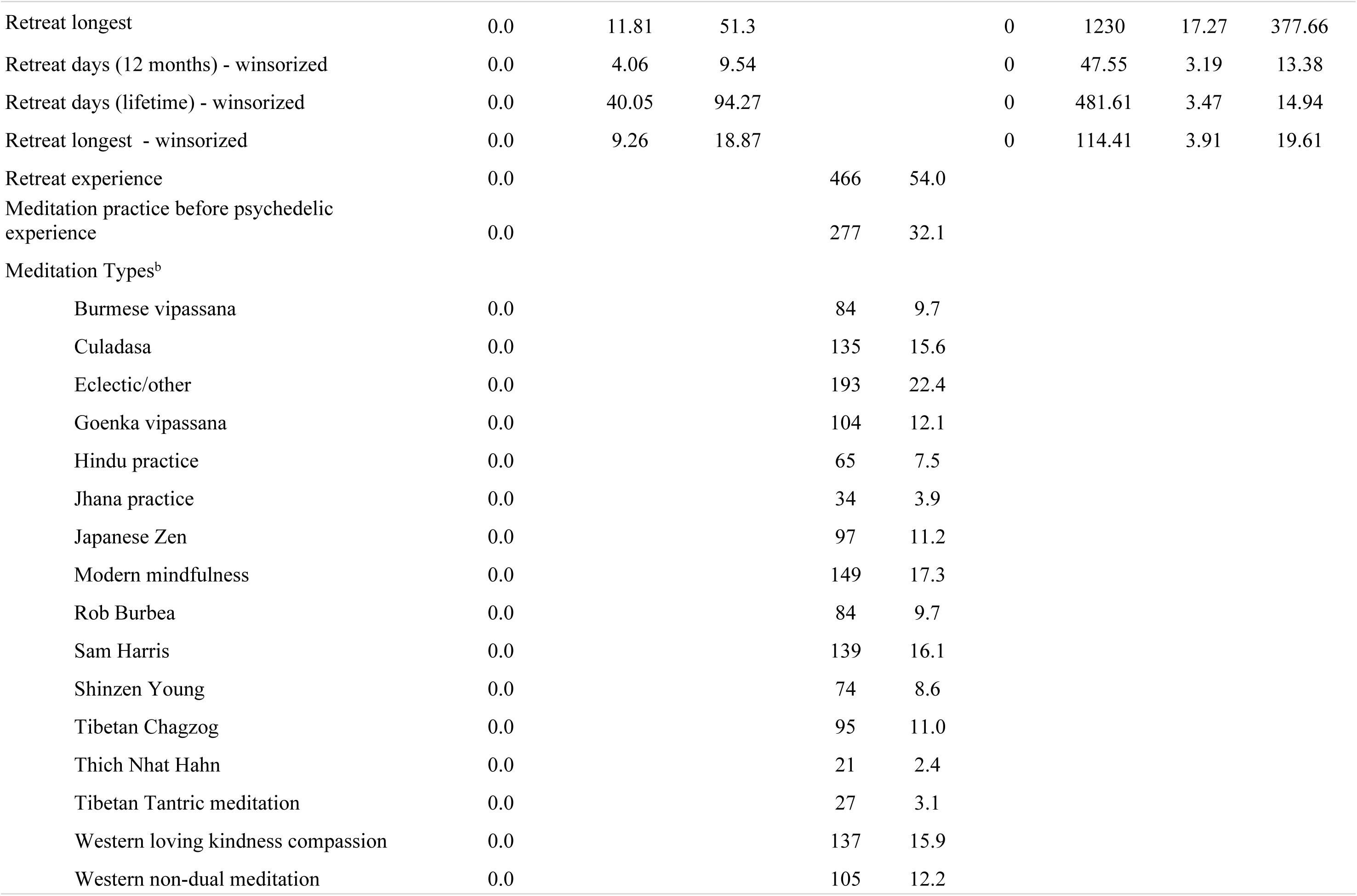
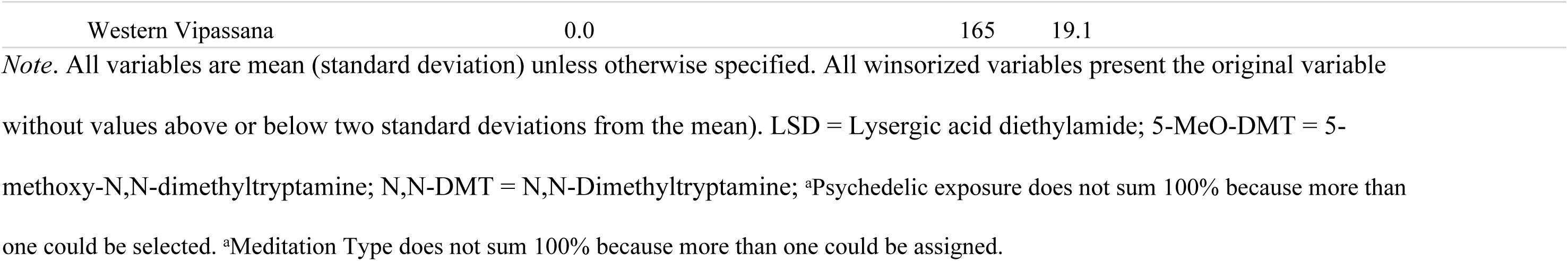
Descriptive Statistics (n = 863)

### Procedure

Data were collected online using Qualtrics. The survey was accessible between 29 October 2020 and 8 June 2021. No financial reward for participation was offered. Before starting the survey, individuals were required to indicate their written informed consent by agreeing to the following statement: ‘It is up to you to decide whether to take part or not; choosing not to take part will not disadvantage you in any way. I have read and understand the explanations, I am at least 18 years old, have a good understanding of the English language, and have maintained a regular meditation practice (at least 3 times per week) during the last 12 months.’ This study received ethical approval from (masked) and was performed in accordance with the 1964 Declaration of Helsinki and its later amendments.

### Measures

Given the absence of standardized self-report measures for capturing the relationship between psychedelic use and meditation practice, we employed a range of self-report questions that were specifically developed for this study. Aiming to capture aspects not yet examined by existing research in this area, the last author (masked) developed an initial pool of survey items, which were then iteratively refined through feedback from and discussions with the third author (masked).

### Outcome Variable

The primary outcome variable was a single-item question that asked participants: “Overall, do you believe that your psychedelic experience(s) have influenced the quality of your regular meditation practice?” Using a 7-point Likert scale, responses ranged from 1 (*strong negative influence*) to 7 (*strong positive influence*). All survey items included are available at the Open Science Framework (https://osf.io/56utj/?view_only=fb526198b6e3492ab90d5a7baffa9ca6).

### Predictor Variables

#### Individual Factors

Self-reported demographic variables included participant age, gender, and education (in years of schooling). Two life satisfaction questions ("Overall, how satisfied are you with your life nowadays?" and "Overall, to what extent do you feel that the things you do in your life are worthwhile?") were drawn from questions asked as part of the UK Census (66). Using an 11-point Likert scale, responses ranged from 0 (*not at all*) to 10 (*completely*).

Personality traits were measured using the Big Five Inventory (BFI; John et al., 1991), which captures the dimensions of openness to experience, conscientiousness, extraversion, agreeableness, and neuroticism using a 5-point Likert scale ranging from 1 (*disagree strongly*) to 5 (*agree strongly*). In this study, Cronbach’s α ranged from 0.74 to 0.87 for these five subscales.

We also captured alcohol, tobacco, and cannabis use. Alcohol use was assessed using an adapted item from screening questions used by the National Institute on Alcohol Abuse and Alcoholism (NIAAA, 2011). Participants were asked about the use of alcohol (“During the last 12 months, how often did you usually have any kind of drink containing alcohol?”) and had ten response options ranging from 0 (*not at all*) to 9 (*everyday*). Tobacco (“During the last 12 months, how often, on average, have you smoked or vaporized tobacco?”) and cannabis (“During the last 12 months, how often, on average, have you smoked or vaporized cannabis/weed?”) were similarly assessed but with five response options ranging from 0 (*not at all*) to 4 (*everyday*).

#### Psychedelic-Related Factors

The frequency of psychedelic use (which excluded microdosing, MDMA/ecstasy, and ketamine) was assessed over the last 12 months and over an individual’s lifetime. Participants were also asked their age when they started consuming psychedelic substances.

The types of psychedelic substances that participants had used at any point in their life were captured by providing a list with the following options: psilocybin/magic mushrooms or truffles, LSD or LSD derivatives, ayahuasca, N,N-DMT, 5-MeO-DMT, mescaline, and others (i.e., an open text box in which participants could specify additional psychedelic substances). Given participants could report exposure to multiple psychedelic substances, binary codes (0 = No, 1 = Yes) were generated for exposure to a given psychedelic substance.

To assess the context and set and setting in which participants used psychedelics, single-item questions were asked about intention setting (“Over your lifetime, when you used psychedelics how often did you set clear intentions before a psychedelic experience?”), attention to safety (“Over your lifetime, when you used psychedelics how often did you pay attention to ensuring the setting was safe and supportive?”), and being surrounded by trusted individuals (“Over your lifetime, when you used psychedelics how often did you pay attention to ensuring you were with people that you trusted?”). Responses ranged from 0 (*never*) to 4 (*always*).

Finally, to understand participants’ experience with psychedelic use, participants were asked about their psychedelic framework (“I have an adequate framework or worldview for understanding or making sense of my psychedelic experience(s)”). Responses ranged from 1 (*disagree strongly*) to 6 (*agree strongly*).

#### Meditation-Related Factors

Frequency of meditation practice (“During the last 12 months, how regularly have you, on average, practiced formal sitting meditation (for at least 20 minutes per session)?”) was assessed with response options ranging from a minimum of 3 (*3 times a week*) to 8 (*twice a day or more*) and was included as a continuous variable. Participants were also asked about their number of years of regular meditation practice (i.e., at least three days a week) and their starting age for practice. To understand participants’ experience with meditation practice, participants were asked about the adequacy of their meditation framework (“I have an adequate framework or worldview for understanding or making sense of my meditation experiences.”). Responses ranged from 1 (*disagree strongly*) to 6 (*agree strongly*).

Participants were also asked whether they had any retreat experience, the number of retreat days in the last twelve months and in their lifetime as well as the length of their longest retreat. Responses were recorded in number of days. Participants were also asked if they first started meditation practice or if they first used a psychedelic.

Finally, to assess meditation practice, participants were invited to qualitatively describe their meditation practice background. Participant responses were then individually coded into 17 practice types by the fourth (masked) and sixth (masked) authors with expertise in modern mindfulness and Buddhist meditation practices. A bottom-up approach was used for coding (inductive derivations of a large number of categories which were then consolidated) while keeping important theoretical considerations in mind for the classification of meditation practices (e.g., Dahl et al., 2015; Lutz et al., 2015). This was done in part due to practical considerations given the varying lengths and specificity of responses from participants. Many participants reported studying under specific contemporary teachers (e.g., Culadasa, Rob Burbea, Sam Harris, Shinzen Young), and in cases where these teachers have created meditation systems that draw on multiple traditions and/or sources in innovative ways that could not be consolidated under a single rubric, the meditation style is coded with the name of the contemporary teacher. Individual names of contemporary teachers were not used when the practices could be consolidated under a general rubric (e.g., Western Non-Dual Meditation). In cases where individuals reported practicing in specific traditional lineages (e.g., Burmese Vipassanā), the practice is coded under the tradition’s name. A significant number of participants (*n* = 193) reported idiosyncratic practices that involved an assorted range of techniques that they themselves invented or were drawn from numerous teachers and traditions. These were coded as “eclectic/other.” The final practice types are listed in the Meditation-Related Factors subsection of Table 1. Response codes ranged from 0 (participant did not mention the practice or teacher) to 1 (participant listed the practice or teacher). A single participant could have reported multiple practice types. For example, one participant responded to the inquiry with “Mindfulness meditation as taught in Sam Harris’ Waking Up app Open awareness practices as taught by Loch Kelly.” In this case, two codes were applied to this response including the teachings of Sam Harris, and Western Non-Dual Meditation. Binary codes (0 = No, 1 = Yes) were created for exposure to each of the 17 categories.

## Data Analysis

### Overview and preliminary analyses

All data, R code, study items, and consent form used in the study are available at the Open Science Framework (https://osf.io/56utj/?view_only=fb526198b6e3492ab90d5a7baffa9ca6). The analyses were planned and conducted following data collection and were not preregistered. While pre-registration may be preferred, it is worth noting that the use of cross-validation combines exploratory and confirmatory methods. Many model configurations are simultaneously explored in training data, but the performance of the best model is subsequently confirmed in new (held-out) data.

For descriptive purposes, we first examined zero-order correlations between the outcome and the predictor variables using Pearson’s correlation coefficient. We report false discovery rate (FDR; Benjamini & Hochberg, 1995) corrected *p*-values for these analyses to control type 1 error rates.

### Machine learning model training and evaluation

Machine learning analyses were conducted using the ‘tidymodels’ ecosystem (69) in R (70). We used machine learning for three reasons. First, when standard (i.e., ordinary least squares; OLS) linear regression is used with high-dimensional, correlated predictors, it often yields high variance models that overfit the training data such that they do not generalize well and therefore perform poorly with new data (64,65). Second, the broader class of linear models (e.g., OLS linear regression, ridge, LASSO, and elastic net regressions) may yield biased models that also perform poorly with new data if the true data-generating process for the outcome is non-linear (e.g., non-linear or interactive effects among predictors and outcome; (71). Third and more generally, when model performance/fit is evaluated with the same data that were used to train the model, the performance will often overestimate how well the model will perform with new data (65). Machine learning methods address the first two issues by simultaneously considering several statistical algorithms that adopt different approaches to address the problems of model bias and variance (due to overfitting) by optimizing the bias-variance trade-off (65,72). Machine learning methods address the third issue by evaluating model performance using new (i.e., held-out) data that were not used to train the models.

In this study, we used three statistical algorithms, OLS linear regression, elastic net linear regression, and random forest. We included OLS linear regression because it is a highly interpretable parametric model that is well-understood and frequently used in the social sciences. We included elastic net linear regression because it is another interpretable parametric model, but it uses regularization techniques that penalize the parameter estimates to yield simpler models that may not overfit training data to the same degree as OLS linear regression (73). We included random forest because it is a flexible, non-parametric model that may outperform linear models when the data-generating process is non-linear (74). It also is often more robust to overfitting than OLS linear regression. Furthermore, both elastic net and random forest often outperform OLS linear regression when multi-collinearity among predictors is present (75).

We fit all models, tuned hyperparameters, and selected the best model configuration using three repeats of 10-fold cross-validation (i.e., 30 held-out folds; Kuhn and Johnson, 2013). For elastic net, we tuned two hyperparameters, lambda (λ; the penalty parameter) and alpha (⍺; the mixing parameter that dictates the proportion of L1 vs L2 penalties). For random forest, we also tuned two hyperparameters, mtry (the number of predictors randomly selected to consider for each split) and minimum node size (the minimum observations needed for additional splitting). We used 1500 trees for all random forest configurations. OLS linear regression does not allow tuning any hyperparameters. Sensible values for each of the hyperparameters for elastic net and random forest were provided by relevant tidymodels functions (from ‘tune’ package) and hyperparameter plots were reviewed to confirm that an adequate range of values was considered. We used mean R^2^ across the 30 held-out folds to tune these hyperparameters, select the best model configuration, and evaluate the performance of that best model.

### Feature engineering

Several feature engineering steps were undertaken to improve model performance. For all three statistical algorithms, we winsorized the predictors to two standard deviations above and below the mean to minimize the influence of outliers (Tukey, 1962). For all algorithms, we also imputed missing data on the predictors using a k-nearest neighbor (KNN) approach implemented within ‘tidymodels’ using package guidelines and function defaults (69). KNN identifies the observations that are most similar to the missing values (i.e., its “nearest neighbors”) and uses the mean value across those cases for imputation (81). We reduced the dimensionality of the predictors somewhat by substituting the first principal component from principal components analyses (PCA) performed with groups of highly correlated predictors that were identified by preliminary exploratory data analysis using the full sample (83). These groups of variables included: 1) two items examining life satisfaction, 2) 12-month and lifetime psychedelic use, and 3) 12-month and lifetime retreat practice. Finally, we applied the Yeo-Johnson power transformation to all quantitative predictors to normalize their distributions for use in the linear statistical algorithms (OLS linear regression and elastic net; 84).

### Feature importance

We planned to use three distinct methods to interpret the best model and identify its important predictors depending on the algorithm that performed best. First, we planned to calculate Shapley Additive Explanations (SHAP; 85) values to assess feature importance associated with each predictor in the best model. Using a concept drawn from cooperative game theory, SHAP values index the average marginal contribution of a given predictor to the predicted outcome for each observation across all possible combinations of predictors (i.e., local SHAPs 86). SHAP values are model agnostic (i.e., can be calculated for any statistical algorithm) and have useful properties including additivity (i.e., values for each observation are computed independently and then summed), symmetry (i.e., average values of two features should be equal if they contribute equally to the overall model), and null contribution (i.e., a feature that does not contribute to the model will have a value of 0). Plots of the relationship between predictor values and local SHAP values can aid in the interpretation of the direction of predictor effects.

Moreover, the global importance (across all observations) for any predictor can be quantified by averaging the absolute value of the local SHAP values across all observations. Larger global SHAP values suggest greater relative importance of a given predictor to the predictive capacity of the model.

Second, we used a feature ablation procedure to assess the incremental contribution of each predictor to the best model. Specifically, for each predictor we compared two nested models, a full model that contained all predictors and a reduced model that removed (i.e., ablated) the relevant predictor. We used an implementation of Bayesian estimation provided by the ‘tidyposterior’ package (89) to estimate the increase in performance for the full model relative to reduced models using R^2^ from the best 30 held-out folds. Predictors are considered important if the posterior probability that R^2^ is greater for the full vs. reduced model exceeds 0.95. Like SHAP, this feature ablation procedure can be used with any statistical algorithm. We sequentially evaluated predictors (ordered by their global SHAP values) until we reached a predictor that did not have a probability > .95 of increasing R^2^ from the reduced model.

Finally, if a linear model (OLS regression or elastic net) was selected as the best-performing algorithm, we planned to review and report its standardized parameter estimates (i.e., *β*s) to rank order the magnitude of the effects of its predictors on the outcome. This third method was not possible for random forest because that algorithm is non-parametric.

## Results

### Descriptive Statistics

Descriptive statistics for all variables are presented in Table 1. The outcome variable, psychedelic experiences improving the quality of regular meditation practice, ranged from 1 (*strong negative influence*) to 7 (*strong positive influence*) and had a mean of 5.49 (*SD* = 1.24). This indicates that, on average, participants reported their meditation quality was improved by psychedelic use (i.e., between “slight positive influence” and “moderate positive influence”). In terms of coded meditation practices, the most frequently mentioned type of practice was eclectic and self-created practices (*n* = 193, 22.0%) followed by Western Vipassana (*n* = 165, 19.1%) and practices taught by Sam Harris (*n* = 139, 16.1%). Number of practices coded for participants ranged from 0-7 with a median of two practices (*M* = 1.98, *SD* = 1.15).

### Correlation Analysis

Correlations between the outcome and each predictor are reported in Supplemental Table 1. Pearson’s correlation coefficient (*r*) values ranged from -.17 to .34 and more than half the predictors (*k* = 28) had a statistically significant association with the outcome even after the FDR correction. The outcome variable had the strongest association with the psychedelic-related variables including psychedelic use (12 months *r* = .34, 95% CI = [.28 to .40], *p* < .001), setting intentions during psychedelic use (*r* = .30, 95% CI = [.23 to .36], *p* < .001), and exposure to N,N-DMT (*r* = .23, 95% CI = [.16 to .29], *p* < .001). Amongst individual factors, the outcome variable had the strongest associations with higher cannabis use (*r* =.18, 95% CI = [.12 to .25], *p* < .001), higher scores on openness to experience (*r* =.16, 95% CI = [.10 to .23], *p* < .001) and higher scores on agreeableness (*r* =.15, 95% CI = [.09 to .22], *p* < .001). Finally, amongst meditation-related variables, the outcome variable had the strongest associations with having less retreat experience (*r* = -.17, 95% CI: -.23 to -.10, *p* < .001) and engaging in eclectic and self-created practices (*r* =.11, 95% CI: .04 to .18, *p* = .004).

### Machine Learning Analysis

#### Best Model Configuration

We used mean R^2^ values across the 30 held-out folds to select the best statistical algorithm and associated hyperparameters. The elastic net linear regression (with λ = 0.19 and ⍺ = 0.20) had the highest mean R^2^ (R^2^ = 0.266, *SE* = 0.013) among the three algorithms. The mean R^2^ was 0.252 (SE = 0.011) and 0.251 (SE = 0.012), respectively, for random forest (with mtry = 5 and minimum node size = 30) and OLS linear regression.

Therefore, all subsequent analyses were conducted with the elastic net linear regression model. Across the 30 held-out folds, the root mean squared error for this model was 1.04, and the mean absolute error was 0.848 (with the outcome measured on a 7-point scale).

#### Feature Importance

As described earlier, we used three complementary methods to examine the feature importance of the predictors in the elastic net model: 1) SHAP values, 2) feature ablation, and 3) standardized coefficients. Global SHAP values for the predictors are presented in Figure 1. This method suggested that the most important features were psychedelic factors: psychedelic use (lifetime/12 month) and setting intention during psychedelic use. The global SHAP values associated with these two predictors (0.25 and 0.15, respectively) were 2 – 4 times greater than any other predictors in the model. The next three important variables were agreeableness (0.07), exposure to N,N-DMT (0.07), and longest retreat (0.07).

**Figure 1.**
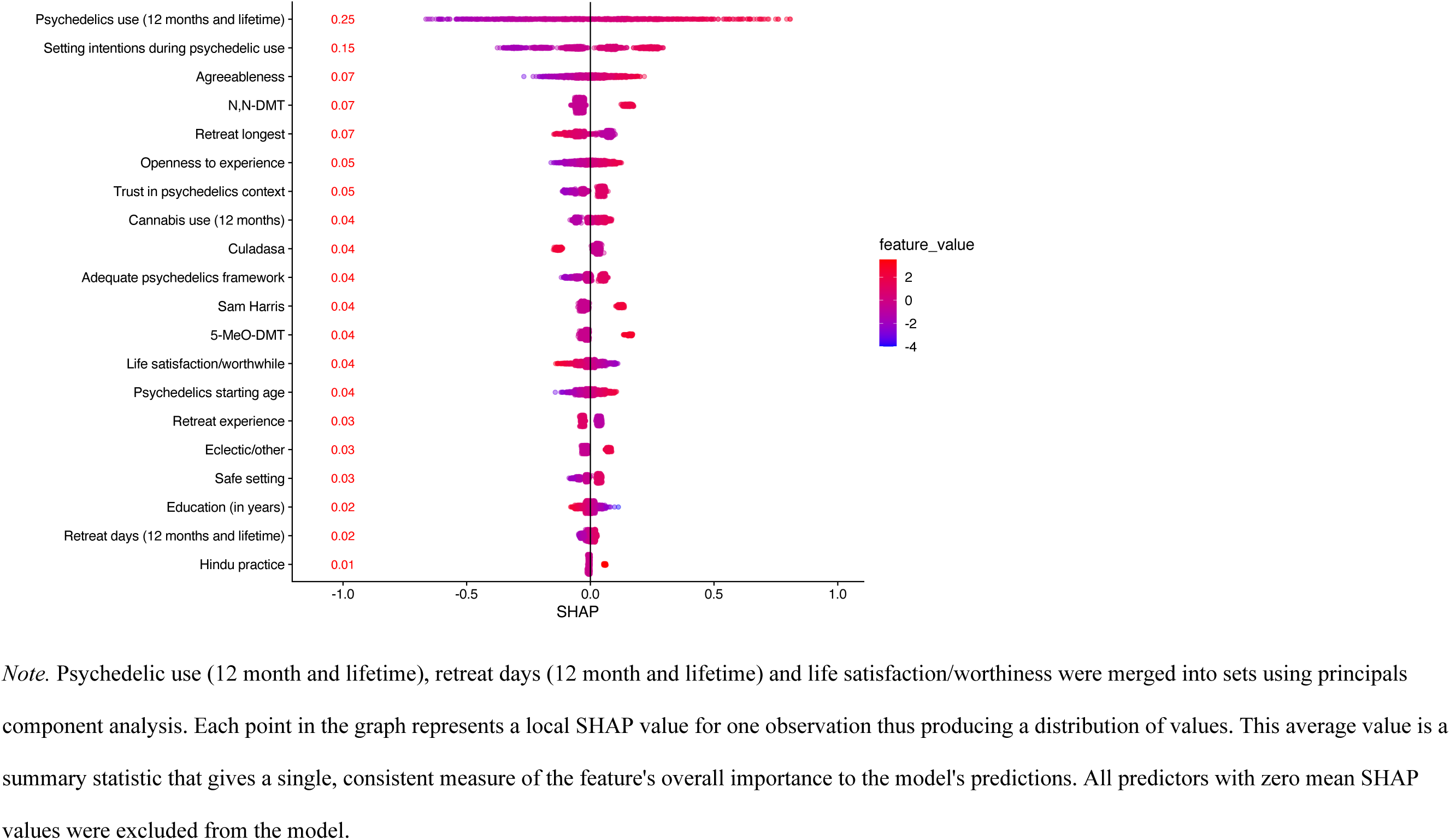
Most Important Variables in the Elastic Net Model organized by the SHAP Values.

We next used feature ablation to sequentially evaluate the incremental contribution of each of the top predictors (ordered by Global SHAP values) to the model. See Table 2 for complete results. For each predictor, we estimated the posterior distribution for the increase in R^2^ when that predictor was added to the reduced model that contained all other predictors. From this distribution we calculated the mean increase in R^2^, the 95% credible interval for this increase, and the posterior probability that this increase was > 0 (i.e., the probability that mean R^2^_full_ mean R^2^_reduced_for that predictor among the held-out folds). Psychedelic use was associated with mean increase in R^2^ of .048 (95% CI = [.032-.064]), with a posterior probability > .999 that this predictor increased model performance. Setting intentions during psychedelic use was associated with a mean increase in R^2^ of .019 (95% CI = [.013-.024]), with a posterior probability > .999 that this predictor increased model performance. Agreeableness was associated with a mean increase in R^2^ of 0.006 (95% CI = [.002, .011]) with a posterior probability of .985 that this predictor increased model performance. N,N-DMT was associated with a mean increase in R^2^ of 0.005 (95% CI = [.001, .009]) with a posterior probability of .970 that this predictor increased model performance. Longest retreat was associated with a mean increase in R^2^ of .001 (95% CI = [-.002, -.002]) and the probability that this predictor increased model performance was low (posterior probability = .553). As the posterior probability was >.95, no further predictors were evaluated.

**Table 2.** Posterior Probabilities of the Full vs. Compact Models.

| **Model** | **Mean R<sup>2</sup>** | **Mean Difference** | **95% CI Lower** | **95% CI Upper** | **Posterior Probability** |
| --- | --- | --- | --- | --- | --- |
| Full Model | .263 |  |  |  |  |
| Compact Model 1 (w/o psychedelic use) | .215 | .048 | .032 | .064 | > .999 |
| Compact Model 2 (w/o setting intentions during psychedelic use) | .246 | .019 | .013 | .024 | > .999 |
| Compact Model 3 (w/o agreeableness) | .257 | .006 | .002 | .011 | .985 |
| Compact Model 2 (w/o N,N-DMT) | .260 | .005 | .001 | .009 | .970 |
| Compact Model 3 (w/o longest Retreat) | .264 | .001 | -.002 | .002 | .553 |

*Note.* CI = credible interval; w/o = without. Predictors are considered important if the posterior probability that R<sup>2</sup> is greater for the
full vs. reduced model exceeds 0.95

To provide a third perspective on feature importance, we display the standardized coefficients for the predictors from the elastic net linear regression in Figure 2. Consistent with the two prior feature importance methods, the four most important predictors, based on the absolute value of their standardized coefficients, were psychedelic use (lifetime/12 month, standardized estimate *β* = 0.30), setting intentions during psychedelic use (*β* = 0.18), agreeableness (*β* = 0.09) and N,N-DMT (*β* = 0.08). See Figure 2 for the list of variables deemend most important.

**Figure 2.**
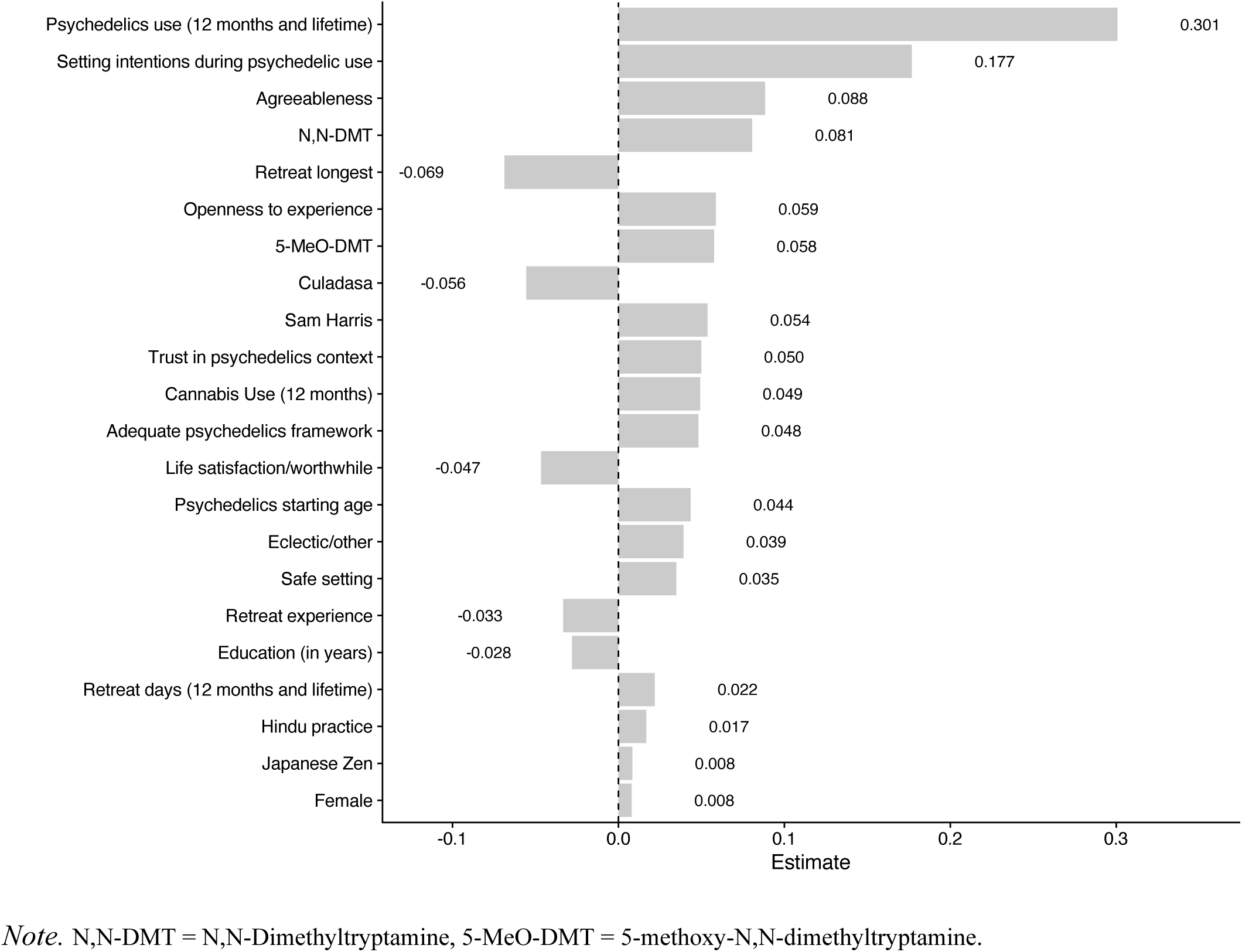
Most Important Variables in the Elastic Net Model Organized by the Value of Standardized Coefficients.

## Discussion

The present study sought to examine which amongst a range of individual, psychedelic-related and meditation-related factors were associated with the perception that psychedelic experiences have positively influenced the quality of one’s meditation practice. This is an important question to understand in the context of the psychedelic renaissance given individuals may be increasingly interested in combining these two modalities. As can be seen from the table of zero-order correlations (Supplemental Table 1), more than half the predictor variables had statistically significant associations with the outcome. As such, we sought to rigorously identify the most important features among this set of predictors. To accomplish this, we tested a range of multivariable machine learning algorithms to find the best-performing model and utilized multiple approaches to assess feature importance (i.e., regression coefficients, SHAP values, feature ablation). Elastic net outperformed linear regression and random forest in the present study.

Across all approaches used to assess feature importance, we found that greater psychedelic use (lifetime/12 month) was the variable most likely to be associated with the perception that psychedelics benefit meditation practice (ΔR^2^ = .048). This is consistent with prior work suggesting that regular use of psychedelics was associated with greater psychological well-being and increased mindfulness practice (20,29,91). The present study extends these findings suggesting that regular use of psychedelics is also associated with greater perceived benefits of psychedelic use on meditation practice for regular meditators. Setting intentions for psychedelic use was also associated with the perception that psychedelic use is beneficial to meditation practice (ΔR^2^ = .019). Prior work suggested that having intentions is predictive of having a peak or mystical experience during psychedelic use (50). Indeed, intention setting – as part of the set and setting hypothesis (i.e., internal and external conditions influence psychedelic experience; 50,51) – is believed to be an important aspect of the subjective theory of how participants perceive psychedelics provide therapeutic benefits (92). The present study builds on this finding suggesting that for psychedelic users, setting intentions may impact not only the acute subjective experience of psychedelic use (50) but may spill over into activities like meditation whose subjective experience may relate to the experience of psychedelic use.

Additionally, we found that two other variables: agreeableness (ΔR^2^ = .006) and exposure to N,N-DMT (ΔR^2^ = .005) were associated with the perception that psychedelics were beneficial to meditation practice, albeit more weakly than psychedelic use and setting intention. The association between agreeableness and the perceived benefit of psychedelic use on meditation practice is in line with prior research indicating that agreeableness may be associated with mystical-type experiences for psychedelic users (61,62) and there is limited evidence to suggest that psychedelic use may enhance agreeableness (93). Interestingly, openness to experience was not found to be an important variable in our cross-sectional analysis. In contrast, in a recent experimental study that administered psilocybin or placebo during a five-day meditation retreat (n=39), openness to experience predicted positively perceived psychedelic experiences (21).

Additionally, exposure to N,N-DMT was also associated with the perception that psychedelics were beneficial to meditation practice. Research on N,N-DMT has found that exposure is associated with subjective experiences of transcendence, unity, and ego dissolution (94,95) and may influence neural activity in ways that are parallel to meditation practice (96).

Focusing on the four variables that were consistently found to be most important across all approaches, a profile emerges of individuals who are most likely to perceive their psychedelic use to benefit meditation practice. These individuals may be those who see psychedelic use as a practice – one that is done regularly and intentionally. They may also be higher on agreeableness and may have exposure to N,N-DMT. Regular and intentional psychedelic use may in theory allow greater opportunities for integration between psychedelic use and meditation practice. It is possible that individuals who engage in regular psychedelic use perceive it as a practice akin to meditation practice and are thereby may be more likely to find compatibility between these two approaches. It has been suggested that compatibility may be a necessary condition to perceive benefits from these co-occurring practices (30). Additionally, the exposure to N,N-DMT, which prior research has suggested may produce more intense and profound experiences relative to other orally administered psychedelics, may also aid individuals in perceiving their psychedelic experience to benefit meditation practice (96–98). It is less clear how agreeableness may contribute to individuals perceiving their psychedelic experience to benefit meditation practice. Prior research has been mixed as one study found that psychedelic use may lead to increases in agreeableness (93) whereas others have found no associations between psychedelic use and agreeableness (12,99). Future longitudinal work is needed to differentiate whether agreeableness is primarily an outcome of the perceived benefits of psychedelics on meditation or a potential causal factor for this association.

Importantly, many other variables showed zero-order correlations with our outcome variable even after an FDR correction and may add further nuance to the picture of those who report their meditation practice benefitting most from psychedelic use. While psychedelic use and intention setting before psychedelic experiences showed some of the largest magnitude associations (*r*s = 0.34 and 0.30, respectively), several other variables may be of interest for exploration in future studies. Factors such as exposure to cannabis use, higher levels of openness to experience, and retreat practice had small but statistically significant positive associations with the perceived benefit of psychedelics on meditation.

Assuming continued movement towards increased access and legalization of psychedelics (100) alongside the continued popularity of meditation (8), there will likely be increased concurrent and simultaneous use of meditation and psychedelics. From a scientific and clinical standpoint, this is largely uncharted territory worthy of exploration. At a broad level, it will be important to develop and test therapeutic models for integrating these strategies. This may include extending prior work investigating the incorporation of psychedelics in established meditation interventions or regular meditation practice (16) and even retreat settings (21) as well as offering meditation as part of the preparation and integration sessions for established psychedelic therapy protocols.

The present findings highlight some specific future directions as well. First, it appears that setting intentions is important and must be considered in any context where meditation and psychedelics are linked. However, it remains unclear how participants set intentions before a psychedelic experience, what these intentions relate to, and which intentions are predictive of a beneficial synergy between psychedelic use and meditation practice (101). Future work in this area could capture the degree of specificity with which intentions are formulated and the domains that intentions are directed toward. Future research could also examine which psychedelics are perceived as most helpful for training particular meditation practices (e.g., mindfulness, compassion, concentration/unification) and explore the differential impact of various meditation-psychedelic combinations on changes in perception. Furthermore, it could be explored whether psychedelics support a deepening of meditation practices by familiarizing meditators with more deconstructed states of consciousness, by making solidified habit patterns and belief structures temporarily more malleable, and by developing a greater capacity to accommodate difficult experiences. Relatedly, it could be pertinent to explore to what extent psychedelic experiences allow meditators to work with psychological material that was previously not accessible or well-integrated through meditation practice alone. Other important factors influencing intention setting could include teacher-student or guide-participant relations, group dynamics of practice communities as well as motivations for psychedelic (102) or meditation use (103).

Another important implication relates to the frequency of psychedelic use whereby greater frequency is associated with greater perceived benefit. This finding could encourage meditators interested in exploring psychedelics, clinicians, and guides to de-emphasize any potentially high expectations brought to a single experience, but rather to consider, as mentioned above, psychedelic use as a longer-term practice akin to psychotherapy or meditation training.

Attitudes traditionally emphasized in both secular (e.g., mindfulness-based stress reduction; 38) and Buddhist meditation traditions (104) may support the adoption of psychedelic use as a complement to meditation. Of particular importance in this regard is an awareness of the potential psychological risks of psychedelics, which remain significant and include challenging experiences, hallucinogen persistent perception disorder, and prolonged psychosis (for a review, see Schlag et al., 2022). Identifying the frequency of psychedelic use that minimizes risks while maximizing the benefits of meditation practice presents a pertinent task for future research.

Further, it remains to be more thoroughly understood which individual characteristics, such as personality traits (or profiles of traits), might predict whether psychedelic use, meditation practice, and a combination of both are perceived as beneficial or, conversely, detrimental and thus considered contraindicated.

### Limitations

Several important limitations must be taken into account when interpreting the results. First, the nature of the cross-sectional design prevents the determination of any cause-and-effect relationship between what we defined as outcome and predictor variables. It is possible that those who perceive the benefits of psychedelic use on meditation are more likely to engage in regular use of psychedelics or intention setting rather than vice versa. It is also possible that the relationships we have found are driven by variables that we have not captured (e.g., worldviews, psychological well-being, mental health, purpose in life, and understanding of appropriate psychedelic use, etc.). Future studies could use longitudinal and experimental approaches to determine causality. It would be particularly illuminating to randomly assign individuals to receive or not receive psychedelics and to examine the impact of meditation practice. Although important work has already begun in this area (e.g., Griffiths et al., 2018; Singer et al., 2024), it would be valuable to examine the effects on a range of non-self-report outcomes measures (e.g., neuroimaging, behavioral tasks) theoretically impacted by some forms of meditation (e.g., attention regulation, prosociality; 42). Second, while the present sample was large, it was a non-representative convenience sample which makes generalization difficult. Similar to a previous online survey of psychedelic users (63), our sample included substantially more male than female participants, which needs to be considered when interpreting the results, particularly given that population-based surveys and other research consistently suggest that female respondents engage in meditation more frequently than male respondents (106–108). Conducting a survey study on a controversial topic such as the use of psychedelics in the context of meditation practice (while psychedelics remain illegal in most places in the world), there is a likelihood for sampling bias (109), because individuals who have either extremely favorable or extremely unfavorable attitudes or experiences might be disproportionately attracted to participate. Future studies will ideally use a more representative sample and may also benefit from oversampling of less representative groups in psychedelic research (i.e., racial/ethnic minority participants; 110). Third, many of the items used in the present study, although based on previous research studies and developed due to the lack of established standardized measures in this area, have not been psychometrically validated. Additionally, the use of a single item to assess the outcome may have introduced unreliability due to the conceptual one-dimensionality of the assessment, potential lack of variability, and error variance (111). Future studies would benefit from the development and use of validated scales. Our survey did not capture whether participants used psychedelics and meditation concurrently or simultaneously, as this may influence their perception of the benefits of psychedelics on meditation. Finally, we utilized a conservative approach (i.e., feature ablation with Bayesian hierarchical linear modeling) to identify a final set of most important features. While we can feel confident in what was found (i.e., low likelihood of type I error), there may have been important variables that were missed (i.e., type II error).

## Data Availability

All data, R code, study items and consent form used in the study are available at the Open Science Framework (https://osf.io/56utj/?view_only=fb526198b6e3492ab90d5a7baffa9ca6)

## Acknowledgments

The salary of SBG was supported by the National Center for Complementary & Integrative Health of the National Institutes of Health under Award Number K23AT010879 (SBG). The salary of OS was supported by Olle Engvist Foundation. The salary of CAW was partially supported by NIMH R01MH116969, NCCIH R01AT011002, the Tommy Fuss Fund and a Young Investigator Grant from the Brain & Behavior Research Foundation.

---

**Supplemental Table 1.** Pearson Correlation Coefficients between the Outcome Variable and each of the Predictors.

| Variable | *r* | 95 % *CI* |  | *p*<sub>raw</sub> | *p*<sub>FDR</sub> |
| --- | --- | --- | --- | --- | --- |
|  |  | *LL* | *UL* |  |  |
| ***Individual Factors*** |  |  |  |  |  |
| Age | -.01 | -.08 | .06 | .771 | .817 |
| Female | .05 | -.02 | .11 | .164 | .217 |
| Education (in years) | -.06 | -.13 | .01 | .077 | .123 |
| Life satisfaction | .13 | .06 | .19 | <.001 | <.001 |
| Life worthwhile | .13 | .07 | .20 | <.001 | <.001 |
| Openness to experience | .16 | .10 | .23 | <.001 | <.001 |
| Neuroticism | -.11 | -.17 | -.04 | .002 | .006 |
| Conscientiousness | .09 | .02 | .15 | .011 | .022 |
| Agreeableness | .15 | .09 | .22 | <.001 | <.001 |
| Extraversion | .07 | .00 | .14 | .042 | .071 |
| Alcohol use (12 months) | -.01 | -.08 | .05 | .668 | .738 |
| Tobacco use (12 months) | .09 | .03 | .16 | .006 | .012 |
| Cannabis use (12 months) | .18 | -.08 | .06 | <.001 | <.001 |
| ***Psychedelic Factors*** |  |  |  |  |  |
| Psychedelics use (12 months) | .34 | .28 | .40 | <.001 | <.001 |
| Psychedelics use (lifetime) | .15 | .08 | .21 | <.001 | <.001 |
| Psychedelics starting age | .10 | .04 | .17 | .002 | .007 |
| Adequate psychedelics framework | .20 | .13 | .26 | <.001 | <.001 |
| Trust in psychedelics context | .19 | .12 | .25 | <.001 | <.001 |
| Setting intentions during psychedelic use | .30 | .23 | .36 | <.001 | <.001 |
| Safe setting | .22 | .16 | .29 | <.001 | <.001 |
| Psilocybin | .06 | .00 | .13 | .064 | .107 |
| LSD | .08 | .02 | .15 | .013 | .025 |
| Ayahuasca | .14 | .07 | .20 | <.001 | <.001 |
| 5-MeO-DMT | .17 | .11 | .24 | <.001 | <.001 |
| N,N-DMT | .23 | .16 | .29 | <.001 | <.001 |
| Mescaline | .09 | .03 | .16 | .005 | .012 |
| Other | .10 | .04 | .17 | .003 | .008 |
| ***Meditation Factors*** |  |  |  |  |  |
| Years of regular practice | -.01 | -.08 | .05 | .695 | .752 |
| Frequency of regular practice | -.05 | -.12 | .02 | .138 | .192 |
| Meditation starting age | -.04 | -.10 | .03 | .279 | .336 |
| Adequate meditation framework | .08 | .01 | .14 | .023 | .044 |
| Retreat experience | -.17 | -.23 | -.10 | <.001 | <.001 |
| Retreat days (12 months) | -.10 | -.17 | -.03 | .003 | .009 |
| Retreat days (lifetime) | -.05 | -.12 | .01 | .128 | .192 |
| Retreat longest | -.07 | -.14 | -.01 | .031 | .056 |
| Meditation practice before psychedelic experience | .002 | -.06 | .07 | .944 | .962 |
| Burmese Vipassana | -.10 | -.16 | -.03 | .004 | .010 |
| Culadasa | -.10 | -.16 | -.03 | .004 | .010 |
| Eclectic/other | .11 | .04 | .18 | .001 | .004 |
| Goenka Vipassana | -.01 | -.07 | .06 | .826 | .858 |
| Hindu practice | .09 | .03 | .16 | .006 | .013 |
| Jhana practice | -.04 | -.10 | .03 | .287 | .338 |
| Japanese Zen | .04 | -.03 | .11 | .231 | .291 |
| Modern mindfulness | .03 | -.03 | .10 | .327 | .377 |
| Rob Burbea | -.06 | -.13 | .01 | .080 | .125 |
| Sam Harris | .07 | .01 | .14 | .034 | .060 |
| Shinzen Young | -.05 | -.12 | .02 | .141 | .192 |
| Tibetan Chagzog | .05 | -.02 | .12 | .141 | .192 |
| Thich Nhat Hahn | .02 | -.05 | .08 | .620 | .700 |
| Tibetan Tantric meditation | -.04 | -.11 | .02 | .199 | .257 |
| Western loving kindness compassion | .00 | -.07 | .07 | .980 | .980 |
| Western non-dual meditation | .04 | -.03 | .11 | .243 | .300 |
| Western Vipassana | -.05 | -.12 | .02 | .137 | .192 |

*Note.* CI = confidence interval, *fdr* = False discovery rate; LSD = Lysergic acid diethylamide; 5-MeO-DMT = 5-methoxy-N,N-dimethyltryptamine N,N-DMT =
N,N-Dimethyltryptamine, .

## Notes

### Competing Interest Statement

OS was a co-founder of Eudelics AB and has once received a small payment from Mindfully Sweden AB for educational content. CAW has received consulting fees from King & Spalding law firm.

### Funding Statement

The author(s) received no specific funding for this work.

### Author Declarations

This study received ethical approval from University College London (Project ID: 10043/004) and was performed in accordance with the 1964 Declaration of Helsinki and its later amendments.

